# Changes in access to Australian disability support benefits during a period of social welfare reform

**DOI:** 10.1101/2020.04.30.20041210

**Authors:** Alex Collie, Luke R Sheehan, Tyler J Lane

**Author notes:** Corresponding Author Email –.

## Abstract

The Disability Support Pension (DSP) is the major Australian government financial benefit program for people of working age with medical conditions and disabilities that restrict work capacity. Between 2012 and 2018 a series of policy reforms sought to restrict the growth in DSP payments and encourage more people with some work capacity to seek employment. We characterise changes in three markers of access to disability financial support over the reform period (1) DSP recipient rates (2) DSP grant (approval) rates and (3) the rate of unemployment benefit receipt in people with impaired work capacity. Results demonstrate a significant reduction in DSP receipt and grant rates, and significant increase in the rate of unemployment benefit receipt in working-age Australians with work disabling medical conditions and disability. These changes were not distributed uniformly. People whose primary medical condition was a musculoskeletal or circulatory system disorder demonstrated greater declines in DSP receipt and grant rates, while there was a more rapid increase in unemployment benefit receipt among people with primary mental health conditions. Some trend changes occur in periods during which new disability assessment and pension eligibility policies were introduced, though our ability to attribute changes to specific policy changes is limited.

**FUNDING AND ACKNOWLEDGMENTS:** This study was supported by an Australian Research Council Future Fellowship awarded to the first author (FT190100218). De-identified, aggregate data for the study was provided by the Australian Government Department of Human Services and Department of Social Services.

## BACKGROUND

Provision of financial benefits to people with medical conditions and disabilities that completely or partially restrict work capacity has been a feature of social assistance programs in industrialised nations since the mid-20^th^ century. These benefits, while typically very modest, provide a critical source of income for people living with disabling conditions. In many OECD nations over the past 40 years there has been substantial growth in the percentage of working age adults receiving disability benefits. From approximately the mid1990’s governments in countries such as Sweden, the Netherlands and the United Kingdom have responded with policy reforms aimed at reducing access to benefits in order to control what has been described as ‘unsustainable growth’ in program expenditure (Burkhauser, Daly, McVicar, & Wilkins, 2014).

In Australia, the primary disability benefit program is the Disability Support Pension (DSP). Like disability benefits in many other industrialised countries, the DSP is designed to provide financial support to people with permanent physical, intellectual or psychiatric impairments that prevent them from engaging fully in employment. Australia’s rate of public spending on disability income support has risen from approximately 1% of GDP during the 1980’s to approximately 2.4% during the 21^st^ century (Organisation for Economic Cooperation and Development (OECD), 2019a). The DSP is now one of the largest programs of Australian government spending. In 2016–17, DSP expenditure was AUD$16.3 billion or 10.6% of all national social security spending, and there were around 760,000 Australians receiving the DSP (Parliamentary Budget Office, 2018), equivalent to 4.7% of the working age population.

Since the turn of the century, Australia has reformed the DSP program periodically to limit growth in expenditure. Policy changes have included, for example, restricting access to those with less than 15 hours work capacity per week from the prior standard of 30 hours (introduced in 2004), changes to the method of assessing job capacity introduced in 2003 and the introduction of external (i.e., non-government) job capacity assessors from 2007 to 2011. From 2012 this reform program has increased in intensity. Successive Australian governments have embarked on a series of changes to DSP eligibility, assessment and compliance processes. These reforms have sought to further restrict the growth in DSP expenditure, reduce the number of new pensions granted and to encourage people to seek employment. This follows a period of rapid growth in expenditure between 2008/9 and 2011/12, driven largely by an increase in the number of new applications being granted (Parliamentary Budget Office, 2018).

Some of the major reforms have included: requiring most applicants to demonstrate that they have actively participated in job seeking or training for a period of 18 months before applying; a program of eligibility reviews for DSP recipients, initially for those aged under 35 years and later for a broader group of recipients; introduction of a new two-stage eligibility assessment procedure for the vast majority of applicants; and introduction of new impairment tables for assessing the degree of work capacity of applicants and during eligibility reviews.

In other nations such practices have been linked with adverse health and employment outcomes. For example, in England a program of re-assessing the work capacity of one million people receiving disability benefits was associated with an increase in suicides, self-reported mental health issues and growth in anti-depressant prescribing (Barr et al., 2016). Policies that restricted access to disability benefits in Denmark and Sweden were linked with significantly increased odds of unemployment among people with moderate and severe medical conditions (Jensen et al., 2019), and the authors of this study concluded that “…the disability benefit reforms seems to have pushed people with both moderate and severe health problems into a life exposed to the economic stress of living with no or temporary and means-tested benefits.”

Reform of the DSP benefit regime may also affect people with some medical conditions and disabilities more than others. Despite impairment assessment guides making allowance for episodic or fluctuating conditions, the requirement to demonstrate that a condition is stable may be challenging for people with conditions that have an unstable symptom course, who may experience periods in which they are severely debilitated and others in which they have greater functional capacity. International studies suggest that where medical opinion on appropriate treatment options for a given condition differs, disability benefit applicants ability to demonstrate that their condition is fully treated may vary according to the experience, training and expertise of the assessing medical practitioner (McAllister & Leeder, 2018). People with psychosocial disabilities may find it more difficult to access benefits under policy regimes that place greater weight on provision of biomedical information to support decision making (McAllister, 2019). Increasing the administrative burden on applicants, through new practices such as requiring applicants to collate and present primary medical evidence, may have a disproportionate impact on people with cognitive, intellectual or psychological conditions (Hong, 2019; Moynihan, Herd, & Harvey, 2014). People with these conditions may have more challenges in gathering complex medical information, completing the required paperwork and communicating their situation to those in positions of influence.

Research on the impact of the recent Australian policy reforms is limited. Government reports suggest that the reforms have reduced the number of people receiving DSP benefits, and that there have been a concurrent increase in the number of people with disability and impaired work capacity receiving the lower rate of unemployment benefit, known as the Newstart Allowance or NSA (Parliamentary Budget Office, 2018). To our knowledge, there is no published data on whether the changes differentially impact people with different disabilities and medical conditions. There has been little analysis on the patterns of change in benefits, nor a detailed characterisation of the relative magnitude of changes in markers of benefit access over this period of reform. Most analysis has been limited to publicly available data on the number of DSP recipients, which may be characterised as lagging indicators of policy reform given the long median duration of benefit receipt and the slow rate of exits from these benefit schemes. Changes to eligibility and application processes are more likely to have short-term impacts on leading indicators such as benefit grant rates. Understanding changes in the other major source of government financial support for people with disability is also important (the NSA), given that the Australian unemployment benefit is among the lowest in the OECD, the level of financial support is much lower than the DSP, and evidence linking unemployment benefit receipt with income poverty and poor health (O’Campo et al., 2015; Roelfs, Shor, Davidson, & Schwartz, 2011).

This study seeks to characterise trends in access to disability support benefits in Australia during the six-year period of policy reform between 2012/13 and 2017/18 financial years. We apply join point analysis to quarterly aggregate benefit data to analyse changes in the rates of DSP recipients, NSA benefit recipients assessed as having partial work capacity, and DSP grants. We analyse data by sex and primary medical condition to assess trends in these outcomes among different groups of recipients. We hypothesised that reductions in disability support during the reform period would be accompanied by an increase in the rate of work disabled people receiving unemployment benefits.

## METHODS

### Data sources and study population

Quarterly data on DSP recipients and NSA recipients with partial capacity (less than 30 hours work capacity per week) were provided by the Australian Government Department of Social Services through a data request for the six year period July 2012 to June 2018. These data represent the number of recipients for each of the benefit programs at the conclusion of each quarter. Data on DSP claims granted were provided by the Australian Government Department of Human Services through a data request for the same time period. These data represent the number of claims approved during each quarter. The time period was selected to correspond with period over which multiple reforms to DSP application, assessment and compliance processes occurred, as described in the introduction.

Data was provided by sex (male, female), age group (16-24, 25-34, 35-44, 45-54, 55-64, 65+) and by the person’s primary medical condition. For the purposes of analyses we grouped medical conditions into six categories that are consistent with Australian Government benefit reporting. These categories also represent the five most common medical condition groups (at July 2018) for DSP recipients and a sixth category which encompasses the remaining medical conditions. Primary medical condition is determined during the eligibility assessment process as the medical condition with the highest impairment rating. The six categories were (1) mental health conditions; (2) musculoskeletal conditions; (3) intellectual or learning disability; (4) nervous system conditions; (5) circulatory system conditions; and (6) all other conditions. Each category includes a number of individual medical conditions, for example the mental health condition category includes major depression, anxiety, schizophrenia, bipolar disorder, post-traumatic stress disorder and a range of other individual psychological or psychiatric conditions. Quarterly data on the resident population of Australia by age and sex for the entire observation period were obtained from the Australian Bureau of Statistics for use as denominators in rate calculations.

### Statistical analyses

We first calculated the count of recipients or grantees on each of the outcomes in total, by sex and primary medical condition in each quarter, as well as the percent change in counts between the first and last quarter of the time series. We then calculated crude rates for each outcome at each quarter in the time series, expressed as the count per 1000 residents of working age (15 to 65 years). To examine outcomes by sex, we calculated the rate per 1000 working age male residents (for males) and the rate per 1000 working age female residents (for females). It was not possible to age-standardise rates as small cell sizes were suppressed in the primary data files to maintain privacy, and this led to a substantial amount of missing age data.

Trends in rates were analysed using join-point regression analysis (Statistical Methodology and Applications Branch, 2019). This method describes changes in time series data trends by connecting several different line segments on a log scale at regression join-points. The method identifies points in a time-based data series where a statistically significant change in the linear slope of the trend occurs. These change points or breaks in the data series are labelled as join-points. The analysis begins by assuming that there are 0 join-points (indicating a straight line) and then tests through an iterative series of statistical comparisons whether there are one or more statistically significant join-points. Model selection used a Monte Carlo permutation test with a maximum of 4499 permutations. The final model for each data series represents the maximum number of statistically significant join-point segments where the probability of an overall type 1 error (alpha) was less than 0.05 (Kim, Fay, Feuer, Midthune, & 2000).

In addition, the quarterly percentage change (QPC) in rates for each join-point line segment were estimated. The QPC is tested to determine if it is statistically different from the null hypothesis that the percent change is 0%. In the final model, each join-point indicates a statistically significant change in trend occurring at a particular time point in the data series. These changes in trends may be either an increase or a decrease, and each of the trends is described by the QPC that is calculated for that join-point segment.

To estimate a summary measure of the trend over the complete study period, the average quarterly percentage change (AQPC) is also computed, with 95% confidence intervals. The AQPC is a weighted average of the individual QPCs, with the weights equivalent to the length of the join-point segments. The QPC and AQPC were calculated for each of the outcomes, and stratified by primary medical condition and sex. For all analyses, a p value less than 0.05 was considered statistically significant.

Calculation of rates were performed in Microsoft Excel. Join-point analyses were performed using the Join-point Regression Software – Desktop Version (Version 4.7.0.0) from the Surveillance Research Program of the US National Cancer Institute (Statistical Methodology and Applications Branch, 2019).

The study was approved by the Monash University Human Research Ethics Committee on 20^th^ February 2019 (Project #18401).

## RESULTS

Results of both descriptive and join-point analyses demonstrate substantial changes in the outcome measures across the study period. Table 1 presents data from the first quarter of the study period (extracted at end September 2012), the last quarter of the study period (extracted at end June 2018) and the percentage change between these two time periods, in order to describe overall changes in absolute numbers of benefit recipients and applicants. This data is not adjusted for changes in population size, and is intended only to provide an indication of the volume of people affected by changes in trends on the three study outcomes, to support interpretation of data.

**Table 1.** Summary of outcomes over time series by study group

| Group | Number (%) at Sept 2012 |  |  | Number (%) at June 2018 |  |  | Percent change over study period |  |  |
| --- | --- | --- | --- | --- | --- | --- | --- | --- | --- |
|  | DSP Recipients | NSA Recipients | DSP Claims Granted | DSP Recipients | NSA Recipients | DSP Claims Granted | DSP Recipients | NSA Recipients | DSP Claims Granted |
| Total | 825,205 (100.0) | 122779 (100.0) | 13508 (100.0) | 756,960 (100.0) | 271421 (100.0) | 8540 (100.0) | -8.3 | 121.0 | -36.8 |
| Males | 443,357 (53.7) | 65049 (53.0) | 7283 (53.9) | 402,901 (53.2) | 136997 (50.5) | 4839 (56.7) | -9.1 | 110.6 | -33.6 |
| Females | 381,848 (46.3) | 57730 (47.0) | 6225 (46.1) | 354,059 (46.8) | 134424 (49.5) | 3701 (43.3) | -7.3 | 132.8 | -40.5 |
| Mental Health Condition | 251,946 (30.5) | 43709 (35.6) | 4273 (31.6) | 257,721 (34.0) | 108771 (40.1) | 2587 (30.3) | 2.3 | 148.9 | -39.5 |
| Musculoskeletal Condition | 222,586 (27.0) | 41429 (33.7) | 2777 (20.6) | 163,973 (21.7) | 88185 (32.5) | 1044 (12.2) | -26.3 | 112.9 | -62.4 |
| Intellectual / Learning Disability | 100,091 (12.1) | 2767 (2.3) | 697 (5.2) | 109,172 (14.4) | 5866 (2.2) | 421 (4.9) | 9.1 | 112.0 | -39.6 |
| Nervous System Condition | 41,465 (5.0) | 3774 (3.1) | 766 (5.7) | 42,462 (5.6) | 8246 (3.0) | 485 (5.7) | 2.4 | 118.5 | -36.7 |
| Circulatory System Condition | 34,450 (4.2) | 7605 (6.2) | 704 (5.2) | 24,832 (3.3) | 13444 (5.0) | 337 (3.9) | -27.9 | 76.8 | -52.1 |
| Other Conditions | 174,667 (21.2) | 23495 (19.1) | 4291 (31.8) | 158,800 (21.0) | 46845 (17.3) | 3666 (42.9) | -9.1 | 99.4 | -14.6 |

Note: DSP = Disability Support Pension; NSA Recipients = Newstart Allowance recipients with partial work capacity. In the data provided a small number of cells (with N cases < 5) were suppressed. In these instances we conservatively assumed 1 case per cell and thus the sum of cases per condition may not equal the total N cases.

There was a reduction in the gross number of DSP recipients over the study period, for both males and females, and for people with musculoskeletal, circulatory system and other conditions. There was an absolute increase in the number of DSP recipients with mental health conditions, intellectual / learning disability and nervous system conditions over the study period. There was substantial growth (>75%) in the number of NSA recipients with impaired work capacity in all groups over the study period. Finally, all groups recorded a reduction in the number of DSP claims granted across the study period. The magnitude of this reduction was largest in the musculoskeletal and circulatory system condition groups, and smallest in the other conditions group.

### Overall trends

Table 2 presents the results of join-point analysis for males, females and the total population. This table presents the join-point line segments for each outcome by each group, including the time period and the QPC statistic for each line segment, as well as the average QPC for the whole study period.

There was a significant reduction in the rate of DSP recipients per 1000 working age population over study period. Significant declines of 0.5% per quarter were observed between September 2012 and June 2013, with the rate declining more rapidly between March 2014 and March 2015 (QPC = -0.7%) and between March 2015 and June 2017 (QPC = -1.2%). The rate of decrease slowed in the final year of the time series. Patterns of change were very similar in females. Males were slightly different and did not show any increase in rates in late 2013 as was evidence in females. The rate of receipt was consistently higher in males throughout the time period (Figure 1a).

**Figure 1.**
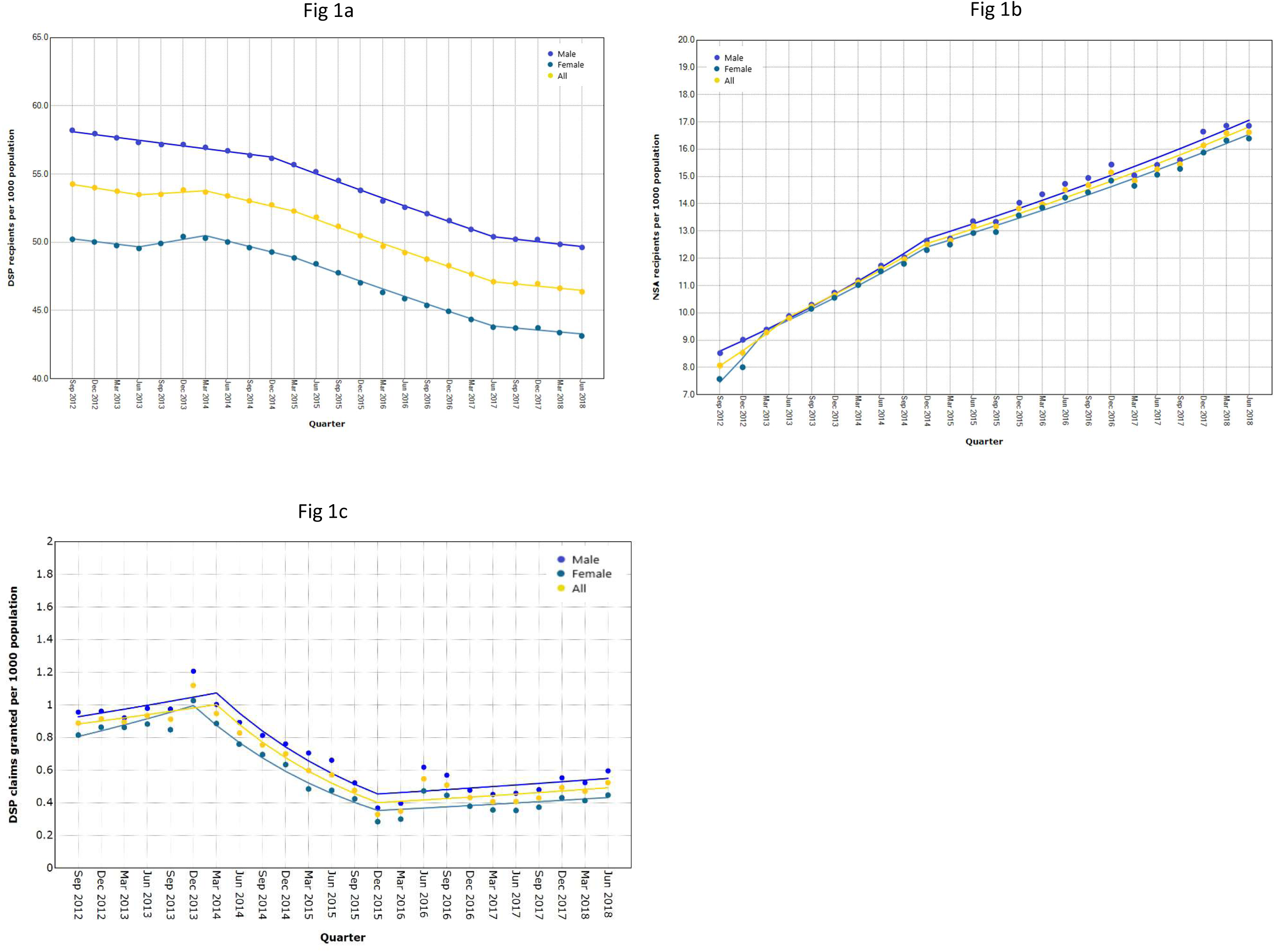
Trends in markers of access to disability income benefits in Australia between July 2012 and June 2018: (1a) Disability Support Pension recipients; (1b) Newstart Allowance recipients with impaired work capacity; (1c) Disability Support Pension claims granted. All data is shown as a rate per 1000 working age population (total: yellow), per 1000 working age males (blue) and per 1000 working age females (green).

**Table 2.** Join-point regression results in total and by sex

| Outcome | Trend 1 |  | Trend 2 |  | Trend 3 |  | Trend 4 |  | Trend 5 |  | Full Range |
| --- | --- | --- | --- | --- | --- | --- | --- | --- | --- | --- | --- |
|  | Period | QPC | Period | QPC | Period | QPC | Period | QPC | Period | QPC | AQPC (95% CI) |
| **DSP Recipient Rate** |  |  |  |  |  |  |  |  |  |  |  |
| Male | Sep 12 – Dec 14 | -0.4* | Dec 14 – Jun 17 | -1.1* | Jun 17 – Jun 18 | -0.4* |  |  |  |  | -0.7* (-0.7, -0.6) |
| Female | Sep 12 – Jun 13 | -0.4* | Jun 13 – Mar 14 | 0.6 | Mar 14 - Mar 15 | -0.8* | Mar 15 – Jun 17 | -1.2* | Jun 17 – Jun 18 | -0.3 | -0.6* (-0.8, -0.5) |
| All | Sep 12 – Jun 13 | -0.5* | Jun 13 – Mar 14 | 0.2 | Mar 14 - Mar 15 | -0.7* | Mar 15 – Jun 17 | -1.2* | Jun 17 – Jun 18 | -0.3* | -0.7* (-0.8, -0.6) |
| **NSA Recipient Rate** |  |  |  |  |  |  |  |  |  |  |  |
| Male | Sep 12 – Dec 14 | 4.4* | Dec 14 – Jun 18 | 2.1* |  |  |  |  |  |  | 3.0* (2.8, 3.2) |
| Female | Sep 12 – Mar 13 | 12.0* | Mar 13 – Dec 14 | 4.1* | Dec 14 – Jun 18 | 2.1* |  |  |  |  | 3.5* (3.2, 3.9) |
| All | Sep 12 – Jun 13 | 7.0* | Jun 13 – Dec 14 | 4.1* | Dec 14 – Jun 18 | 2.1 |  |  |  |  | 3.3* (2.9, 3.7) |
| **DSP Claims Granted** |  |  |  |  |  |  |  |  |  |  |  |
| Male | Sep 12 – Mar 14 | 2.1 | Mar 14 – Dec 15 | -12.3* | Dec 15 – Jun 18 | 2.1 |  |  |  |  | -2.2 (-4.5, 0.0) |
| Female | Sep 12 – Dec 13 | 4.3 | Dec 13 – Dec 15 | -12.1* | Dec 15 – Jun 18 | 2.0 |  |  |  |  | -2.7* (-4.7, -0.6) |
| All | Sep 12 – Mar 14 | 2.1 | Mar 14 – Dec 15 | -12.3* | Dec 15 – Jun 18 | 2.1 |  |  |  |  | -2.5* (-4.6, -0.3) |

QPC = Quarterly Percent Change; AQPC = Average Quarterly Percent Change; DSP = Disability Support Pension; NSA = Newstart Allowance with impaired work capacity; CI = Confidence Interval; \*p<0.05

There was a significant increase in the rate of NSA recipients with impaired work capacity over the study period (AQPC = 3.3%). The rate increased most rapidly between September 2012 and June 2013 (QPC = 7.0%), and continued to increase significantly but at a slower rate between June 2013 and June 2018. Patterns of change were similar between males and females, though the growth during the first segment in females was more rapid and occurred over a shorter time period (Figure 1b and Table 2).

There was a significant reduction in the rate of DSP claims granted over the study period (AQPC = -2.5%). During the period between Mar 2014 and Dec 2015 the rate of decline was 12.3% per quarter. Non-significant trend increases were observed in both males and females for the periods before Dec 2013 and for the final 2 ½ years of the study period between December 2015 to June 2018. Grant rates were substantially lower at the conclusion of the study period than at the beginning, and the patterns were very similar between males and females (Figure 1c).

### Trends by primary medical condition

For all outcomes, changes in trend varied substantially by primary medical condition group (Table 3). Five of the six groups displayed a significant reduction in the rate of DSP recipients over the study period, with the exception being the group of recipients with Intellectual / Learning Disability in which the rate increased by 0.1% per quarter on average. The greatest change in rate was observed in the musculoskeletal condition (AQPC = -1.6%) and Circulatory system condition (AQPC = -1.7%) groups. In these groups the most rapid declines occurred between March 2015 and June 2017. During that same time period the rate of DSP recipients declined significantly across all medical condition groups, though the changes were more modest in the Intellectual / Learning Disability, Mental health condition and Nervous system condition groups. The trend for these three groups also displayed an increase over the first 2 years of the time series, which was statistically significant in most cases. In contrast the musculoskeletal disorders, circulatory system and other condition groups recorded a significant decline in rate over the first year of the time series.

**Table 3.**
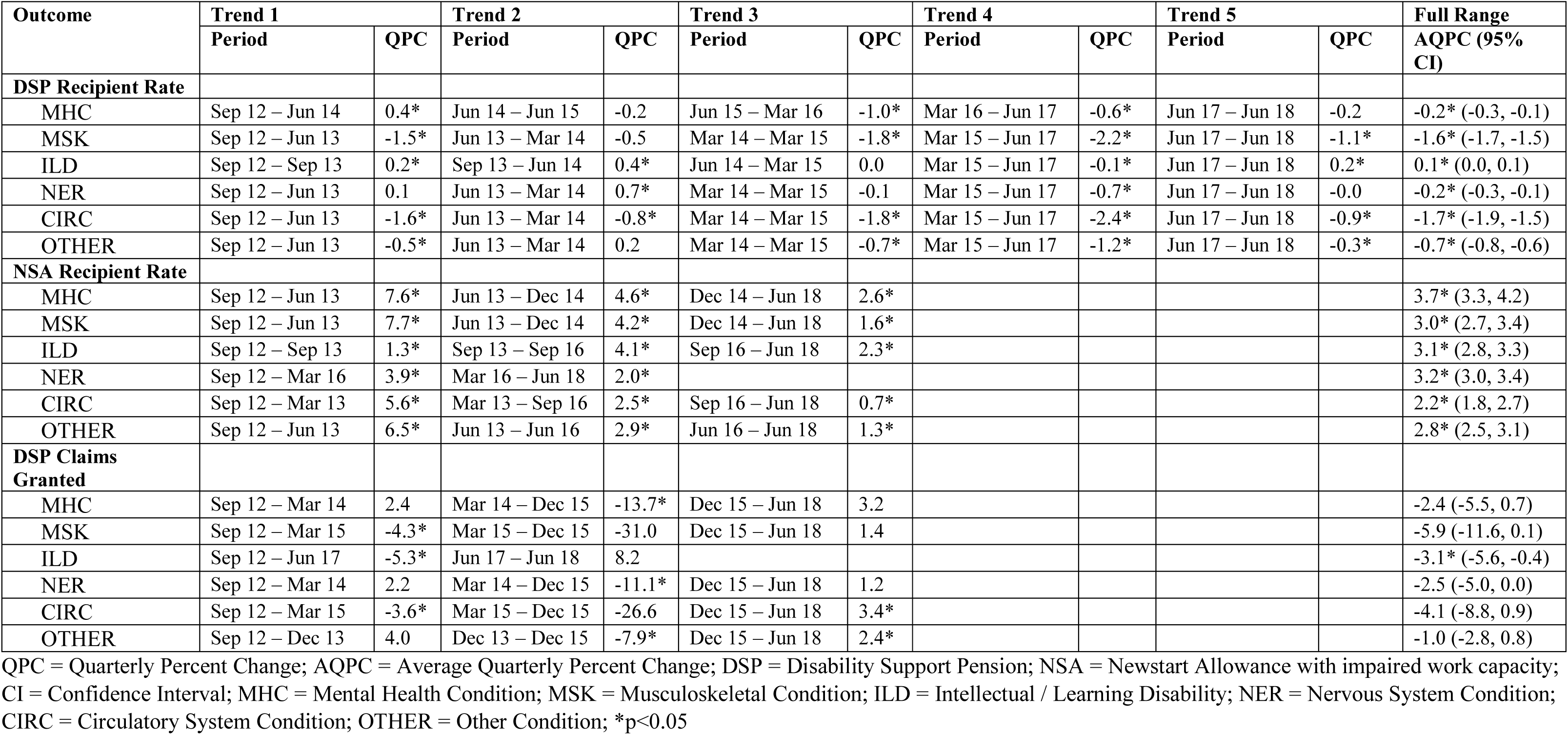
Join-point regression results by primary medical condition

| Outcome | Trend 1 |  | Trend 2 |  | Trend 3 |  | Trend 4 |  | Trend 5 |  | Full Range |
| --- | --- | --- | --- | --- | --- | --- | --- | --- | --- | --- | --- |
|  | Period | QPC | Period | QPC | Period | QPC | Period | QPC | Period | QPC | AQPC (95% CI) |
| **DSP Recipient Rate** |  |  |  |  |  |  |  |  |  |  |  |
| MHC | Sep 12 – Jun 14 | 0.4* | Jun 14 – Jun 15 | -0.2 | Jun 15 – Mar 16 | -1.0* | Mar 16 – Jun 17 | -0.6* | Jun 17 – Jun 18 | -0.2 | -0.2* (-0.3, -0.1) |
| MSK | Sep 12 – Jun 13 | -1.5* | Jun 13 – Mar 14 | -0.5 | Mar 14 – Mar 15 | -1.8* | Mar 15 – Jun 17 | -2.2* | Jun 17 – Jun 18 | -1.1* | -1.6* (-1.7, -1.5) |
| ILD | Sep 12 – Sep 13 | 0.2* | Sep 13 – Jun 14 | 0.4* | Jun 14 – Mar 15 | 0.0 | Mar 15 – Jun 17 | -0.1* | Jun 17 – Jun 18 | 0.2* | 0.1* (0.0, 0.1) |
| NER | Sep 12 – Jun 13 | 0.1 | Jun 13 – Mar 14 | 0.7* | Mar 14 – Mar 15 | -0.1 | Mar 15 – Jun 17 | -0.7* | Jun 17 – Jun 18 | -0.0 | -0.2* (-0.3, -0.1) |
| CIRC | Sep 12 – Jun 13 | -1.6* | Jun 13 – Mar 14 | -0.8* | Mar 14 – Mar 15 | -1.8* | Mar 15 – Jun 17 | -2.4* | Jun 17 – Jun 18 | -0.9* | -1.7* (-1.9, -1.5) |
| OTHER | Sep 12 – Jun 13 | -0.5* | Jun 13 – Mar 14 | 0.2 | Mar 14 – Mar 15 | -0.7* | Mar 15 – Jun 17 | -1.2* | Jun 17 – Jun 18 | -0.3* | -0.7* (-0.8, -0.6) |
| **NSA Recipient Rate** |  |  |  |  |  |  |  |  |  |  |  |
| MHC | Sep 12 – Jun 13 | 7.6* | Jun 13 – Dec 14 | 4.6* | Dec 14 – Jun 18 | 2.6* |  |  |  |  | 3.7* (3.3, 4.2) |
| MSK | Sep 12 – Jun 13 | 7.7* | Jun 13 – Dec 14 | 4.2* | Dec 14 – Jun 18 | 1.6* |  |  |  |  | 3.0* (2.7, 3.4) |
| ILD | Sep 12 – Sep 13 | 1.3* | Sep 13 – Sep 16 | 4.1* | Sep 16 – Jun 18 | 2.3* |  |  |  |  | 3.1* (2.8, 3.3) |
| NER | Sep 12 – Mar 16 | 3.9* | Mar 16 – Jun 18 | 2.0* |  |  |  |  |  |  | 3.2* (3.0, 3.4) |
| CIRC | Sep 12 – Mar 13 | 5.6* | Mar 13 – Sep 16 | 2.5* | Sep 16 – Jun 18 | 0.7* |  |  |  |  | 2.2* (1.8, 2.7) |
| OTHER | Sep 12 – Jun 13 | 6.5* | Jun 13 – Jun 16 | 2.9* | Jun 16 – Jun 18 | 1.3* |  |  |  |  | 2.8* (2.5, 3.1) |
| **DSP Claims Granted** |  |  |  |  |  |  |  |  |  |  |  |
| MHC | Sep 12 – Mar 14 | 2.4 | Mar 14 – Dec 15 | -13.7* | Dec 15 – Jun 18 | 3.2 |  |  |  |  | -2.4 (-5.5, 0.7) |
| MSK | Sep 12 – Mar 15 | -4.3* | Mar 15 – Dec 15 | -31.0 | Dec 15 – Jun 18 | 1.4 |  |  |  |  | -5.9 (-11.6, 0.1) |
| ILD | Sep 12 – Jun 17 | -5.3* | Jun 17 – Jun 18 | 8.2 |  |  |  |  |  |  | -3.1* (-5.6, -0.4) |
| NER | Sep 12 – Mar 14 | 2.2 | Mar 14 – Dec 15 | -11.1* | Dec 15 – Jun 18 | 1.2 |  |  |  |  | -2.5 (-5.0, 0.0) |
| CIRC | Sep 12 – Mar 15 | -3.6* | Mar 15 – Dec 15 | -26.6 | Dec 15 – Jun 18 | 3.4* |  |  |  |  | -4.1 (-8.8, 0.9) |
| OTHER | Sep 12 – Dec 13 | 4.0 | Dec 13 – Dec 15 | -7.9* | Dec 15 – Jun 18 | 2.4* |  |  |  |  | -1.0 (-2.8, 0.8) |

QPC = Quarterly Percent Change; AQPC = Average Quarterly Percent Change; DSP = Disability Support Pension; NSA = Newstart Allowance with impaired work capacity; CI = Confidence Interval; MHC = Mental Health Condition; MSK = Musculoskeletal Condition; ILD = Intellectual / Learning Disability; NER = Nervous System Condition; CIRC = Circulatory System Condition; OTHER = Other Condition; \*p<0.05

**Figure 2.**
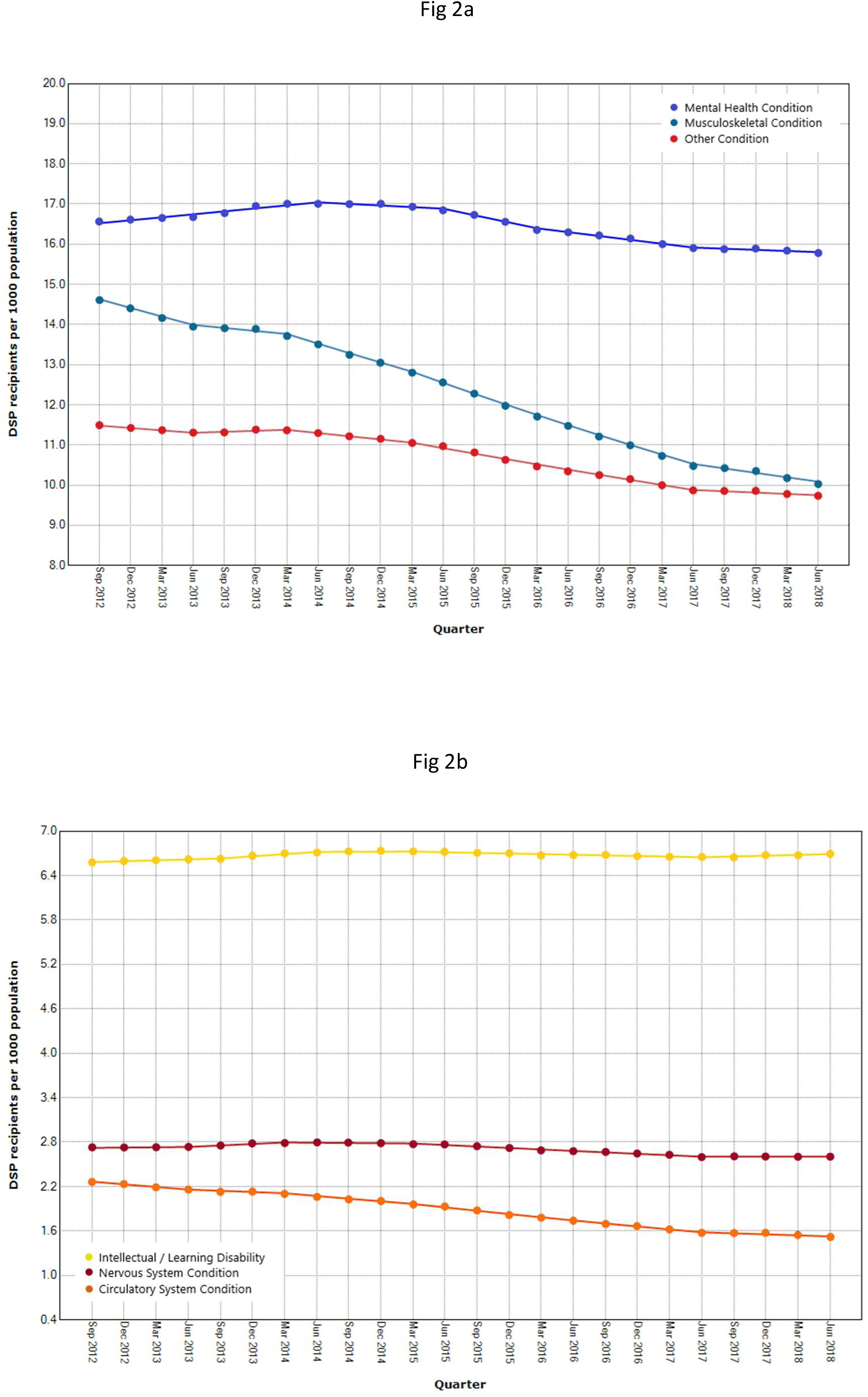
Trends in the rate of Disability Support Pension recipients per 1000 working age population by the recipient’s primary medical condition; (2a) Mental health conditions, Musculoskeletal conditions and Other conditions; (2b) Intellectual / Learning Disability, Nervous system conditions and Circulatory system conditions.

In contrast with DSP recipient rates, the rate of NSA recipients with impaired work capacity increased over the study period across all six groups. The greatest increase in rate was for people with mental health conditions. In this group there was an absolute increase of 149% (more than 65,000 extra recipients) during the study period. This corresponded to an AQPC of 3.7% over the study period. The trend in this group occurred in three distinct segments: an initial rapid increase of 7.6% per quarter between September 2012 and June 2013 followed by a slower but still statistically significant increase of 4.6% per quarter between June 2013 and December 2014. In the final three and a half years of the time series between December 2014 and June 2018 the rate increased at 2.6% per quarter. The pattern of an initial rapid increase in the rate of NSA recipients with impaired capacity followed by a slower increase or stabilisation was observed for the musculoskeletal, nervous system, circulatory system and other condition groups. A different pattern was observed in people with intellectual and learning disability, in whom a gradual increase between September 2012 and September 2013 was followed by a more rapid increase in rate over the rest of the study period.

**Figure 3.**
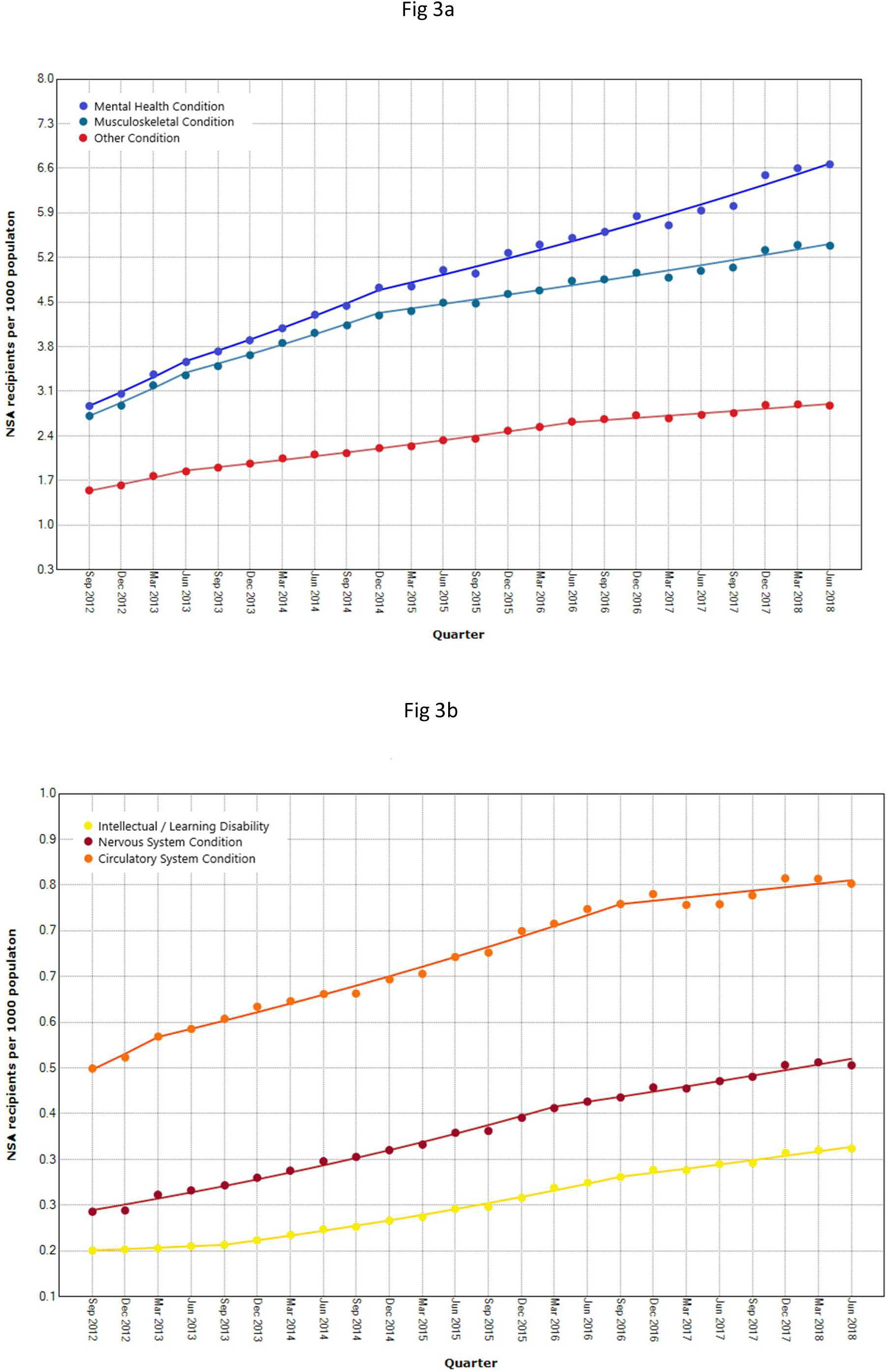
Trends in the rate of Newstart Allowance recipients with impaired work capacity per 1000 working age population by the recipient’s primary medical condition; (3a) Mental health conditions, Musculoskeletal conditions and Other conditions; (3b) Intellectual / Learning Disability, Nervous system conditions and Circulatory system conditions.

Reductions in DSP grant rates were observed in all six medical condition groups, with multiple trend periods reaching statistical significance. The overall reduction in rate was greatest in people with musculoskeletal conditions (AQPC = 5.9%) followed by those with circulatory system conditions (AQPC = 4.1%). The slowest overall reduction was observed in people with other conditions (AQPC = 1.0%). With the exception of people with intellectual and learning disability, all groups displayed a distinct three-phase pattern. In the mental health conditions, nervous system and other condition groups, this was characterised by an initial increase in grant rates over the first six to seven quarters of the time series, then a rapid reduction in grant rates to December 2015, followed by a gradual increase in trend until June 2018. In the musculoskeletal and circulatory system groups, there was an initial reduction until March 2015, followed by a rapid reduction to December 2015, and then as with the other groups a gradual increase in trend until June 2018. The rate of reduction during the middle of the three phases was rapid at 31.0% per quarter in the musculoskeletal condition group, 26.6% per quarter in the circulatory system group, 13.7% per quarter in the mental health condition group and 11.1% in the Nervous system condition group. For all six groups grant rates peaked in December 2013 and reached their lowest point in December 2015 or March 2016. Although not always indicated in the join-point analyses, the most rapid declines in grant rates across all groups were between these two time points.

**Figure 4.**
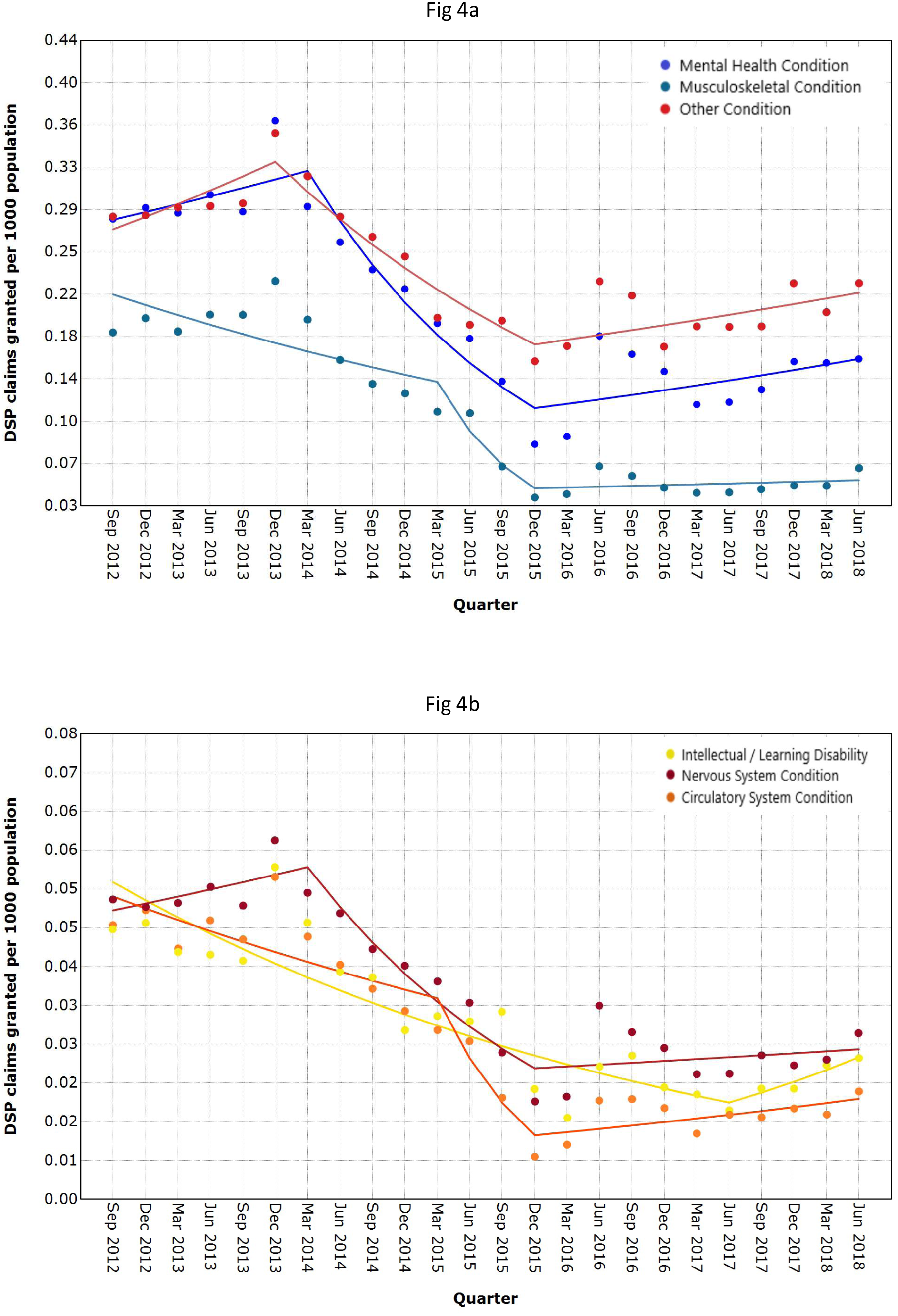
Trends in the rate of Disability Support Pension applications granted per 1000 working age population by the recipient’s primary medical condition; (4a) Mental health conditions, Musculoskeletal conditions and Other conditions; (4b) Intellectual / Learning Disability, Nervous system conditions and Circulatory system conditions.

## DISCUSSION

This study observed significant declines in the number of people accessing disability income support in Australia during a period in which multiple reforms to DSP eligibility criteria, application processes and compliance procedures were implemented. One objective of these reforms was to slow growth in disability benefit receipt. We observed an overall decreasing trend in access to the DSP, characterised both by a reduction in the rate of recipients but also reductions in the rate of applications granted. This was accompanied by a significant increase in the rate of people with disability and impaired work capacity accessing the main unemployment benefit in Australia, the Newstart Allowance. This is both consistent with our hypothesis and with the proposition that restricting access to one working age benefit (i.e, the DSP) has resulted in movement of work disabled people to an alternative benefit (i.e., the NSA).

These changes were not uniformly distributed across the recipient and applicant populations. Trends varied according to the recorded primary medical condition. People with a primary musculoskeletal condition, who historically have been the largest group of DSP recipients (Parliamentary Budget Office, 2018), exhibited the greatest reduction in the rate of DSP claims granted and the second greatest reduction in the rate of DSP recipients. In contrast people with mental health conditions who recently became the largest group of DSP recipients by volume (Parliamentary Budget Office, 2018) had the greatest increase in the rate of NSA recipients with impaired work capacity, and a decrease in the rate of DSP recipients that was slower than the overall trend. By volume, there were nearly 60,000 fewer DSP recipients with a primary musculoskeletal condition at the end of the study period than at the beginning, whereas there were approximately 6,000 more DSP recipients with primary mental health conditions at the end of the study than at the beginning (Table 1).

Changes to benefit and grant rates were not uniform across the study period. There were periods in which changes in outcomes accelerated or slowed. In some instances, periods of more rapid change corresponded with the introduction of specific policy and practice changes. For example, we observed a rapid decline in DSP grant rates during the 2014 and 2015 calendar years. This period overlaps with the introduction of changes to DSP eligibility assessment practices, the intended impact of which was to reduce the inflow (grant rate) to the DSP and thus over time to reduce the number of people receiving disability support benefits. Other major reforms introduced during this period included tightening of the requirements for some DSP applicants to demonstrate that they had been actively participating in a program of support (i.e., job finding, education or training), including for example addition of the criteria that the program of support must be wholly or partially delivered by a government-funded provider. The reduction in grant rates over this period was largest in people with musculoskeletal and circulatory system conditions. One explanation for this finding of differential impact by primary medical condition is that people with musculoskeletal and circulatory conditions are more likely to be assessed as having some work capacity, and thus less likely to be exempted from these assessment and job finding requirements on the basis of condition severity.

Many of the reforms enacted over this period are targeted towards people who do not have a ‘severe impairment’ as measured using the Australian Government Impairment Tables for the Assessment of Work-Related Impairment (Commonwealth of Australia, 2019). For example, compulsory participation in a program of support does not apply to DSP applicants assessed as meeting the criteria for a severe impairment. A severe impairment exists if an individual is judged to have a severe or extreme impairment in a single domain of function, but not if an individual is judged to have moderate or mild impairments across multiple domains of function. Some of the impairment tables include specific guidance as to the use of validated functional assessment instruments by specialist qualified healthcare practitioners, such as the table for assessing intellectual function. Other impairment tables require more subjective judgments of function and do not specify the expertise of assessors. These variations in the rigour and objectivity of functional assessment may partially explain the apparent differential impact of policy reforms across groups with different medical conditions.

One theme of the Australian disability benefit policy reforms since 2012 has been a shifting of the burden of information provision and compliance to the benefit applicant or recipient, and away from the social welfare agency Centrelink. For example, the changes to medical assessment procedures in 2015 required DSP applicants to collect and provide original medical records along with their claim. Applicants are provided with a checklist of the types of medical evidence that they may wish to supply, such as hospital records or x-rays. Under the previous assessment process, the person claiming the DSP was issued with a medical report form to be completed by their treating doctor.

This additional administrative burden may have a particularly adverse effect on people living with disability. A recent analysis found that people who receive support to complete their DSP application claim form are 20% more likely to have their claim granted than those who complete the form without assistance (Hong, 2019). This demonstrates both the importance of information provision and support, but also the challenges for people with disability in completing a long, complex form filled with technical language. The impact of administrative burdens are not equally distributed across society (Herd & Moynihan, 2019). People with less ‘human capital’ (such as lower education, less money, smaller social networks, fewer psychological resources, poorer health or greater disability) will be more negatively impacted by burdens, and will have less access to resources that may help overcome administrative burdens (Herd & Moynihan, 2019).

Some of our findings extend understanding of how people with some highly prevalent conditions interact with social assistance schemes. A prior study of mental illness in Australia reported that while the prevalence of probable common mental disorders had not changed over the period 2001 to 2014, the number of people with mental health conditions accessing the DSP had grown by 51% (Harvey et al., 2017). The present study shows both that this trend of growth in DSP recipients with mental health conditions has reversed in recent years, and that instead there has been a significant growth in the number and rate of people with mental health conditions receiving the NSA. The rate of growth in NSA receipt was faster for people with mental health conditions (3.7% per quarter) than for the other condition categories studied. Australian government spending on unemployment cash benefits sits below the OECD average at 0.65% of GDP (Organisation for Economic Cooperation and Development (OECD), 2019b). The base rate for a single person receiving the NSA is $277.85 per week (as at August 2019) compared with $463.10 per week for a single person receiving the full rate of the DSP. Purely from a public health perspective, this is a concerning trend given evidence that life expectancy is inversely related to the generosity of welfare regimes (Beckfield & Bambra, 2016), and that poverty is a barrier to recovery from mental illness (Weich & Lewis, 1998).

Musculoskeletal conditions account for 23% of the national non-fatal burden of disease in Australia (Australian Institute of Health and Welfare, 2016), with most of this burden borne by people in middle and late working age (35 to 65 years). Four of the top ten causes of nonfatal burden of disease are musculoskeletal diseases, and these include back pain, osteoarthritis, rheumatoid arthritis and other musculoskeletal conditions (Australian Institute of Health and Welfare, 2017). These conditions have multiple contributors including a range of physical, psychological, social, occupational and lifestyle factors and can be challenging to diagnose and treat (Hartvigsen et al., 2018). Our findings demonstrate a significant reduction in the rate of access to social assistance financial benefits among working age Australians with disabling musculoskeletal conditions between 2012 and 2018. The proportion of DSP recipients with a primary musculoskeletal condition fell from 27.0% to 21.7% (Table 1) equivalent to a population standardised rate reduction of 1.6% per quarter (Table 3). The grant rate for people with musculoskeletal conditions fell more sharply, from 20.6% of all claims granted in the first quarter of our time series to 12.2% in the final quarter (−5.9% average QPC). These reductions exceeded those observed for most other conditions, and suggest that people with complex, symptom-based medical conditions such as musculoskeletal conditions may have been disproportionately affected by the recent reforms to the Australian welfare system.

Strengths of this study include the use of population-level data and use of a statistical approach that has been validated in multiple previous studies of the prevalence of health indicators. Our application of this technique to social assistance data is relatively novel. Use of multiple indicators provided a more nuanced understanding of how access to disability benefits changes in response to policy intervention. Some changes were large and observed over short time scales (e.g., changes to grant rates) while others were slower changes over longer time periods (e.g., changes to DSP recipient rates). Examining outcomes by primary medical condition enabled differentiation of trends by an important characteristic, and implications to be drawn regarding the interaction between condition / disability and benefit access. Limitations include that we are unable to directly attribute observed changes in trends to specific policy or practice interventions as would be possible with quasi-experimental methods such as interrupted time series. Being based on aggregate data, the join-point approach also relies on a limited number of observations and thus in smaller samples displays more variation in estimates, making it more difficult to detect meaningful patterns. This was evident in our analysis by medical condition, in which seemingly large changes in trend were non-significant. The join-point approach is however, a valuable tool for examining time-based changes in trends where there have been multiple major reforms implemented at different time periods, as is the case in the Australian social security system over the study period. Data was provided in a way that limited ability to age adjust rates, and thus we report crude rates adjusted for changes in the working age population.

### Conclusions

Between 2012 and 2018 there were large and statistically significant changes in the rate of access to disability support and unemployment benefits for working age Australians with medical conditions and disability that restrict work capacity, in addition to a significant reduction in approval of new disability support pensions. Many more Australians with work disabling conditions now receive unemployment benefits, while significantly fewer receive the higher rate of cash benefits distributed via the disability support pension. These changes were not distributed uniformly. People whose primary condition was a musculoskeletal or circulatory system disorder demonstrated greater changes in DSP receipt and grant rates than those with other conditions, while people with a primary mental health condition demonstrated a more rapid increase in receipt of unemployment benefits than those with other conditions. The changes in trends are also time-varying. Some changes occur in periods during which new disability assessment and pension eligibility policies were introduced, although our ability to attribute changes to specific policy changes is limited. Overall, findings indicate that recent policy reform in the Australian social assistance scheme has had a significant impact on access to the two major government programs of financial support for working age people with serious medical condition and disabilities.

## Data Availability

De-identified, aggregate data for the study was provided by the Australian Government Department of Human Services and Department of Social Services. Data is available from those departments by request. The authors are not able to provide data to any third parties.

